# SARS-CoV-2 Seroprevalence in a University Community: A Longitudinal Study of the Impact of Student Return to Campus on Infection Risk Among Community Members

**DOI:** 10.1101/2021.02.17.21251942

**Authors:** Callum R.K. Arnold, Sreenidhi Srinivasan, Sophie Rodriguez, Natalie Rydzak, Catherine M. Herzog, Abhinay Gontu, Nita Bharti, Meg Small, Connie J. Rogers, Margeaux M. Schade, Suresh V Kuchipudi, Vivek Kapur, Andrew Read, Matthew J. Ferrari

## Abstract

**Background:** Returning university students represent large-scale, transient demographic shifts and a potential source of transmission to adjacent communities during the COVID-19 pandemic.

**Methods:** In this prospective longitudinal cohort study, we tested for IgG antibodies against SARS-CoV-2 in a non-random cohort of residents living in Centre County prior to the Fall 2020 term at the Pennsylvania State University and following the conclusion of the Fall 2020 term. We also report the seroprevalence in a non-random cohort of students collected at the end of the Fall 2020 term.

**Results:** Of 1313 community participants, 42 (3.2%) were positive for SARS-CoV-2 IgG antibodies at their first visit between 07 August and 02 October 2020. Of 684 student participants who returned to campus for fall instruction, 208 (30.4%) were positive for SARS-CoV-2 antibodies between 26 October and 21 December. 96 (7.3%) community participants returned a positive IgG antibody result by 19 February. Only contact with known SARS-CoV-2-positive individuals and attendance at small gatherings (20-50 individuals) were significant predictors of detecting IgG antibodies among returning students (aOR, 95% CI: 3.1, 2.07-4.64; 1.52, 1.03-2.24; respectively).

**Conclusions:** Despite high seroprevalence observed within the student population, seroprevalence in a longitudinal cohort of community residents was low and stable from before student arrival for the Fall 2020 term to after student departure. The study implies that heterogeneity in SARS-CoV-2 transmission can occur in geographically coincident populations.

**Author’s summary:** Despite high seroprevalence observed within the student population, seroprevalence in a longitudinal cohort of community residents remained low and stable from before student arrival for the Fall term to after their departure, implying limited transmission between these subpopulations.

## Background

Demographic shifts, high population densities, and population mobility are known to impact the spread of infectious diseases [1–5]. While this has been well characterized at large scales [6–8], it has proved more challenging to demonstrate at smaller geographic scales [9–11]. The return of college and university students to in-person and hybrid (in-person and online) instruction in the Fall 2020 term during the COVID-19 pandemic represented a massive demographic shift in many communities in the United States (US); specifically, increased population and proportion living in high density living facilities, with a concomitant increase in person-to-person interactions [12]. This shift had the potential to increase SARS-CoV-2 transmission in returning students and to surrounding communities, particularly for non-urban campuses where incidence lagged larger population centers [13]. Modeling analyses conducted prior to student return raised concerns that university re-opening would result in significant SARS-CoV-2 transmission in both the returning student and community resident populations [14,15].

During the Fall 2020 term, many universities in the US experienced high rates of COVID-19 cases among students [16], with a 56% increase in incidence among counties home to large colleges or universities relative to matched counties without such institutions [12]. While there is strong evidence of high incidence rates associated with a return to campus at US colleges and universities [12], the increase in risk in surrounding communities, and transmission rate from campuses to communities, have been less well characterized. The observed increases in COVID-19 cases in these communities cannot be explicitly attributed to campus origin, absent detailed contact tracing.

This investigation reports the results of a longitudinal serosurvey of community residents in Centre County, Pennsylvania, USA, which is home to The Pennsylvania State University (PSU), University Park (UP) campus. The return of approximately 35,000 students to the UP campus in August 2020 represented a nearly 20% increase in the county population [17]. During the Fall 2020 term, more than 4,500 cases of SARS-CoV-2 infections were detected among the student population [18]. Between 7 August and 2 October 2020 (before and just after student return), we enrolled a cohort of community residents and tested serum for the presence of anti-Spike Receptor Binding Domain (S/RBD) IgG, which would indicate prior SARS-CoV-2 exposure [19]. This was repeated in the same cohort during December 2020 (post-departure of students), and we present seroprevalence for both sampling waves. Additionally, returning students were enrolled in a longitudinal cohort, and IgG seroprevalence results are presented from the first wave of sampling (between October and November 2020, prior to the end of the term). The hypothesis tested was that the influx of students during the Fall 2020 term would be correlated with increased SARS-CoV-2 community prevalence.

## Methods

### Design, Setting, and Participants

This human subjects research was conducted with PSU Institutional Review Board approval and in accordance with the Declaration of Helsinki. The study uses a longitudinal cohort design, with two separate cohorts: community residents and returning students. We report on measures from the first two clinic visits for the community resident cohort and the first clinic visit for the returning student cohort.

To assist with recruitment into studies under the Data4Action (D4A) Centre County COVID Cohort Study umbrella, a REDCap survey was distributed to residents of Centre County where respondents could indicate interest in future study participating and provide demographic data. Returning students received a similar survey and were also recruited through cold-emails and word-of-mouth.

Individuals were eligible for participation in the community resident cohort if they were: ≥18 years old, residing in Centre County at the time of recruitment (June through September 2020); expecting to reside in Centre County until June 2021; fluent in English; and capable of providing their own consent. PSU students who remained in Centre County through spring and summer university closure were eligible for inclusion in the community resident cohort as they experienced similar geographic COVID-19 risks as community residents. Participants were eligible for inclusion in the returning student cohort if they were: ≥18 years old; fluent in English; capable of providing their own consent; residing in Centre County at the time of recruitment (October 2020); officially enrolled as PSU UP students for the Fall 2020 term; and intended to be living in Centre County through April 2021. In both cohorts, individuals were invited to participate in the survey-only portion of the study if they were: lactating, pregnant, or intended to become pregnant in the next 12 months; unable to wear a mask for the clinic visit; demonstrated acute COVID-19 symptoms within the previous 14 days; or reported a health condition that made them uncomfortable with participating in the clinic visit.

Upon enrollment, returning students were supplied with a REDCap survey to examine socio-behavioural phenomena, such as attendance at gatherings and adherence to non-pharmaceutical interventions, in addition to their travel history and contact with individuals who were known or suspected of being positive for SARS-CoV-2. Community residents received similar surveys with questions relating to potential SARS-CoV-2 household exposures. All eligible participants were scheduled for a clinical visit at each time interval where blood samples were collected.

### Outcomes

The primary outcome was the presence of S/RBD IgG antibodies, measured using an indirect isotype-specific (IgG) screening ELISA developed at PSU [20]. Further details in the Supplement. The presence of anti-SARS-CoV-2 antibodies has been documented in prior seroprevalence studies as a method of quantifying cumulative exposure [21–23].

### Statistical Methods

Community resident and returning student cohorts’ seroprevalence are presented with binomial 95% confidence intervals. We estimated each subgroup’s true prevalence, accounting for imperfect sensitivity and specificity of the IgG assay, using the **prevalence** package in R. We calculated a 95% binomial confidence interval for test sensitivity of the IgG assay for detecting prior self-reported positive tests in the returning student cohort (students had high access to testing from a common University provider) with a uniform prior distribution between these limits. Prevalence estimates were calculated across possible values of specificity between 0.85 and 0.99. Estimates were not corrected for demographics as participants were not enrolled using a probability-based sample. We assessed demographic characteristics of the tested participants relative to all study participants to illustrate potential selection biases.

Missing values were deemed “Missing At Random” and imputed, as described in the Supplement. We estimated the adjusted odds ratios (aOR) of IgG positivity in the student subgroup using multivariable logistic regression implemented with the **mice** and **finalfit** packages, and two-sided Chi-squared tests for raw odds ratios (OR), and present 95% confidence intervals. We considered the following variables *a priori* to be potential risk factors as they increase contact with individuals outside of a participants’ household [24–27]: close proximity (6 feet or less) to an individual who tested positive for SARS-CoV-2; close proximity to an individual showing key COVID-19 symptoms (fever, cough, shortness of breath); attendance at a small gathering (20-50 people) in the past 3 months; attendance at a medium gathering (51-1000 people) in the past 3 months; lives in University housing; ate in a restaurant in the past 7 days; ate in a dining hall in the past 7 days; only ate in their room/apartment in the past 7 days; travelled in the 3 months prior to returning to campus; and travelled since returning to campus for the Fall term.

We estimated the aOR of IgG positivity at either time point in the community subgroup, with the following risk factors determined *a priori*: being a PSU employee; and the amount of contact with PSU students when “Stay at home” orders are not in place (self-reported on a scale of 1-10). BIC and AIC were used to evaluate the contribution of the variables to the model.

All statistical analyses were conducted using R version 4.1.1 (2021-08-10), with a pipeline created using the **targets** package.

## Results

A total of 9299 community residents were identified through an initial REDCap survey that collected eligibility, demographic, and contact information. 1531 were eligible, indicated willingness to participate, and were enrolled. 1462 completed a first clinic visit between 07 August and 02 October 2020, and 1313 of those completed a second clinic visit between 30 November and 19 February 2020 and for whom both visit 1 and visit 2 samples were analysed. 1410 returning students were recruited using volunteer sampling and 725 enrolled; of these, 684 completed clinic visits for serum collection between 26 October and 21 December 2020.

Among participants with serum samples: the median age community residents was 47 years (IQR: 36-58), with 86.5% between the ages 18-65 years, and for the returning students the median age was 20 years (IQR: 19-21), with 99.7% between the ages 18-65 years; 66.9% of the community residents identified as female and 32.3% as male; 64.5% of the returning students identified as female and 34.6% as male; 92.9% of the community residents identified as white, as did 81.9% of the students. Similar proportions were seen in those enrolled without samples, and among the initial survey respondents (Table 1; Table 2). Although all county residents were eligible for participation, 74.9% of community resident participants were from the 5 townships (College, Ferguson, Harris, Half Moon, Patton) and 1 borough (State College) that form the “Centre Region” and account for ∼59% of Centre County’s population [17] (Figure 1). The median household income group in the community residents providing samples was $100,000 to $149,999 USD (IQR: $50,000 to $74,999; $150,000 to $199,999). The median household income in the county is $60,403 [17]. 47.4% of the county is female, 87.9% white, and 70.3% are between the ages of 18-65 years old [17]. The study cohort is moderately older and more affluent (in part because of the exclusion of returning students), and disproportionately female compared to the general Centre County population.

**Table 1:**
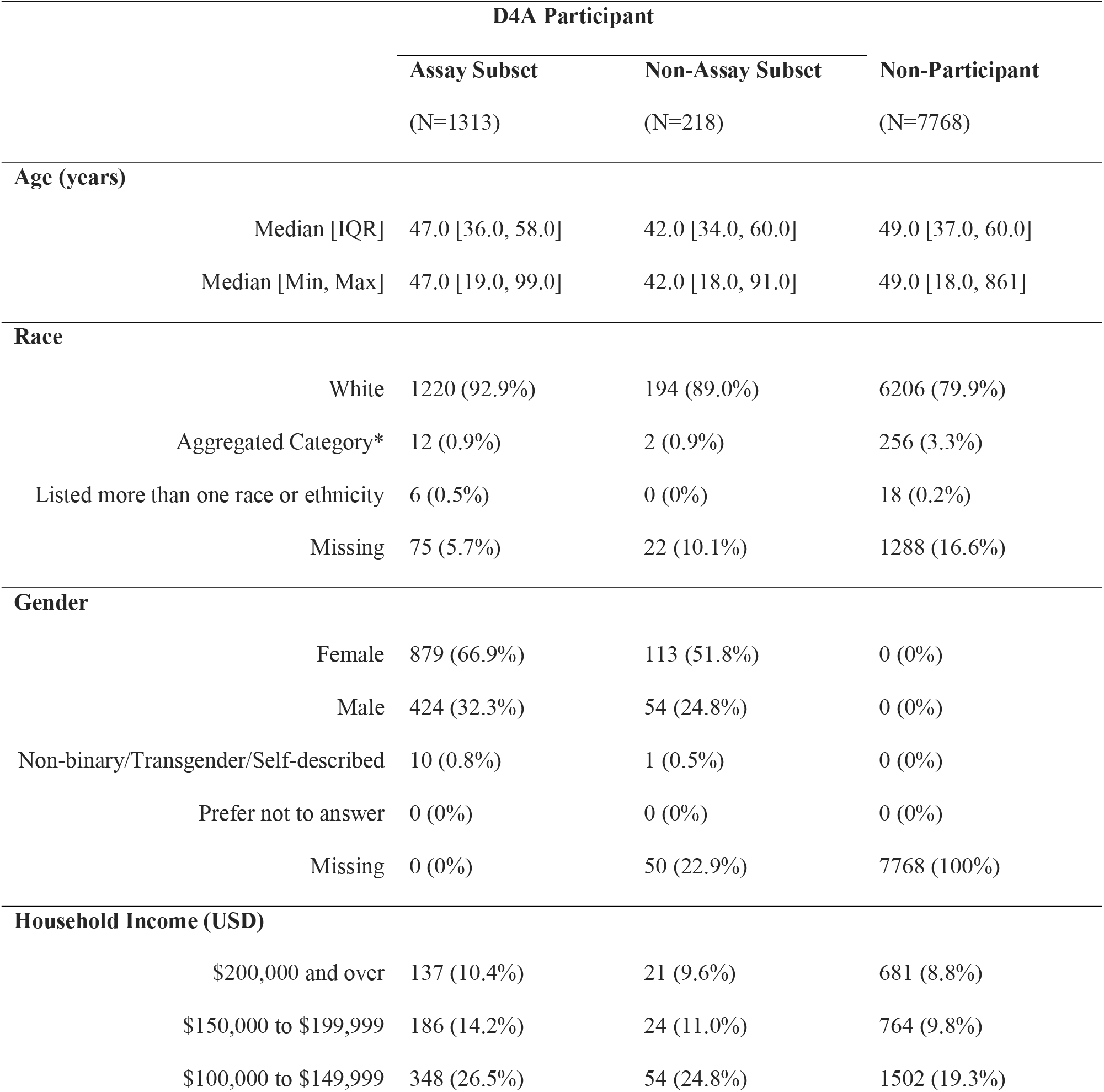
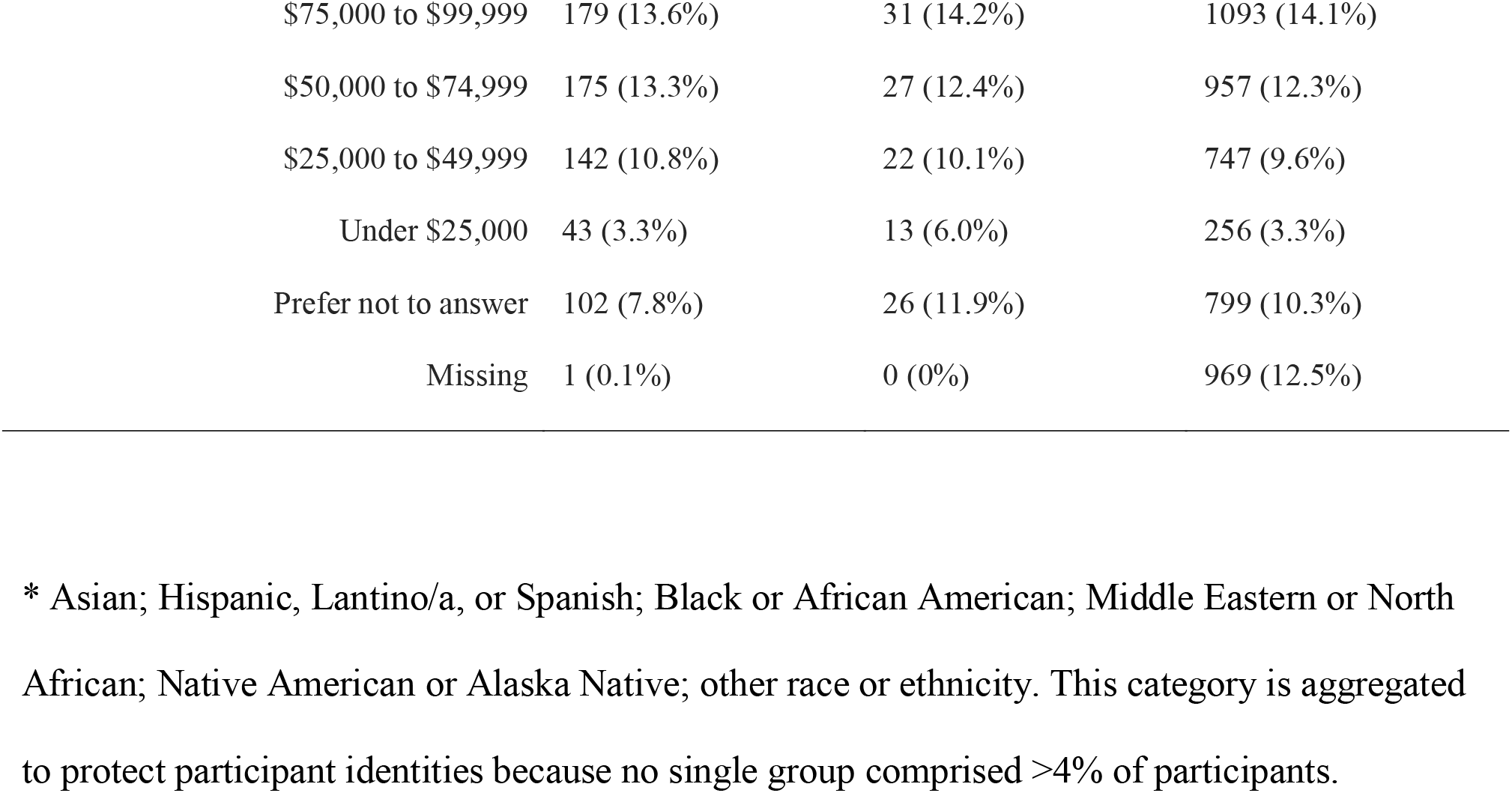
Demographic characteristics of study participants. Non-D4A participants are all participants in the initial anonymous survey from which Data4Action participants were drawn. D4A participants are divided into subsets for which antibody assays were conducted (N=1313) and those for which assays were not conducted (N=218).

|  |  | D4A Participant |  |  |
| --- | --- | --- | --- | --- |
|  |  | Assay Subset | Non-Assay Subset | Non-Participant |
|  |  | (N=1313) | (N=218) | (N=7768) |
| Age (years) |  |  |  |  |
|  | Median [IQR] | 47.0 [36.0, 58.0] | 42.0 [34.0, 60.0] | 49.0 [37.0, 60.0] |
|  | Median [Min, Max] | 47.0 [19.0, 99.0] | 42.0 [18.0, 91.0] | 49.0 [18.0, 861] |
| Race |  |  |  |  |
|  | White | 1220 (92.9%) | 194 (89.0%) | 6206 (79.9%) |
|  | Aggregated Category* | 12 (0.9%) | 2 (0.9%) | 256 (3.3%) |
|  | Listed more than one race or ethnicity | 6 (0.5%) | 0 (0%) | 18 (0.2%) |
|  | Missing | 75 (5.7%) | 22 (10.1%) | 1288 (16.6%) |
| Gender |  |  |  |  |
|  | Female | 879 (66.9%) | 113 (51.8%) | 0 (0%) |
|  | Male | 424 (32.3%) | 54 (24.8%) | 0 (0%) |
|  | Non-binary/Transgender/Self-described | 10 (0.8%) | 1 (0.5%) | 0 (0%) |
|  | Prefer not to answer | 0 (0%) | 0 (0%) | 0 (0%) |
|  | Missing | 0 (0%) | 50 (22.9%) | 7768 (100%) |
| Household Income (USD) |  |  |  |  |
| | \$200,000 and over | 137 (10.4%) | 21 (9.6%) | 681 (8.8%) |
| | \$150,000 to \$199,999 | 186 (14.2%) | 24 (11.0%) | 764 (9.8%) |
| | \$100,000 to \$149,999 | 348 (26.5%) | 54 (24.8%) | 1502 (19.3%) |

|  |  |  |  |
| --- | --- | --- | --- |
| \$75,000 to \$99,999 | 179 (13.6%) | 31 (14.2%) | 1093 (14.1%) |
| \$50,000 to \$74,999 | 175 (13.3%) | 27 (12.4%) | 957 (12.3%) |
| \$25,000 to \$49,999 | 142 (10.8%) | 22 (10.1%) | 747 (9.6%) |
| Under \$25,000 | 43 (3.3%) | 13 (6.0%) | 256 (3.3%) |
| Prefer not to answer | 102 (7.8%) | 26 (11.9%) | 799 (10.3%) |
| Missing | 1 (0.1%) | 0 (0%) | 969 (12.5%) |

\* Asian; Hispanic, Lantino/a, or Spanish; Black or African American; Middle Eastern or North African; Native American or Alaska Native; other race or ethnicity. This category is aggregated to protect participant identities because no single group comprised >4% of participants.

**Table 2:** Demographic characteristics of the returning student participants.

|  | **Assay Subset** | **Non-Assay Subset** |
| --- | --- | --- |
|  | (N=684) | (N=41) |
| **Age (years)** |  |  |
| Median [IQR] | 20.0 [19.0, 21.0] | 20.0 [20.0, 21.0] |
| Median [Min, Max] | 20.0 [18.0, 67.0] | 20.0 [18.0, 32.0] |
| Missing | 1 (0.1%) | 18 (43.9%) |
| **Race** |  |  |
| White | 560 (81.9%) | 27 (65.9%) |
| Aggregated Category* | 86 (12.6%) | 5 (12.2%) |
| Listed more than one race | 32 (4.7%) | 2 (4.9%) |
| Missing | 6 (0.9%) | 7 (17.1%) |
| **Gender** |  |  |
| Female | 441 (64.5%) | 19 (46.3%) |
| Male | 237 (34.6%) | 22 (53.7%) |
| Genderqueer/nonconforming/transgender/different identity | 5 (0.7%) | 0 (0%) |
| Missing | 1 (0.1%) | 0 (0%) |
| **University Housing** |  |  |
| Not Uni housing | 501 (73.2%) | 27 (65.9%) |
| Uni housing | 181 (26.5%) | 8 (19.5%) |
| Missing | 2 (0.3%) | 6 (14.6%) |

\* Asian; Hispanic, Lantino/a, or Spanish; Black or African American; Middle Eastern or North African; Native American or Alaska Native; other race or ethnicity. This category is aggregated to protect participant identities because no single group comprised >4% of participants.

**Figure 1:**
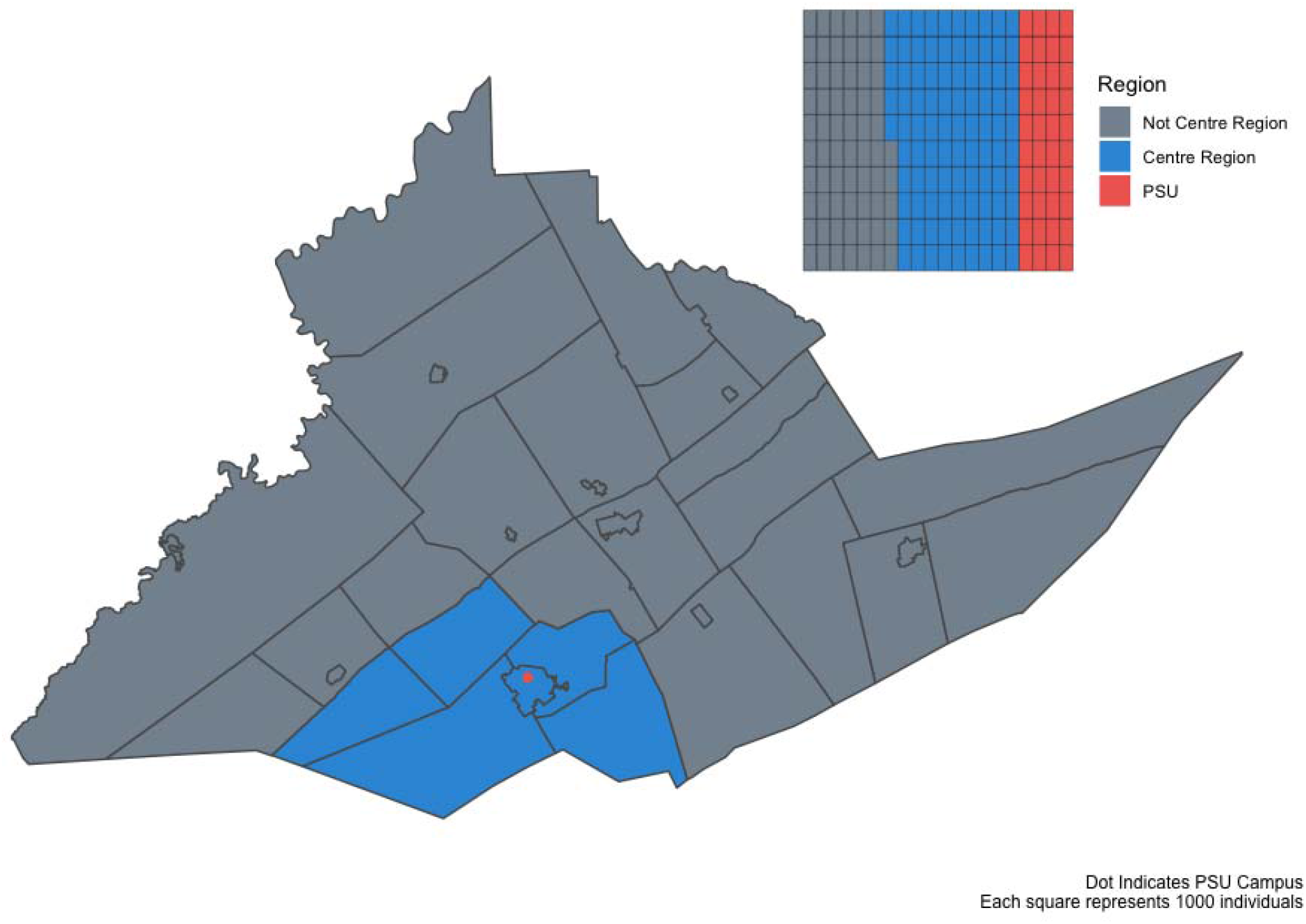
Map of Centre County, Pennsylvania, USA. Blue indicates the 5 townships and 1 borough that comprise the Centre Region. Red indicates the location of The Pennsylvania State University (PSU), University Park (UP) Campus. Inset illustrates the proportion of the county population in each region; PSU indicates the estimated student population that returned to campus for the Fall 2020 term.

Of the returning student participants, 673 (92.8%) had at least one test prior to enrollment in the study; of these, 107 (15.9%) self-reported a positive result (Table 3). Of these, 100 (93.5%) indicated that this test result occurred after their return to campus (median: 25 September; IQR: 10 September, 07 October). Of the 684 returning students with an ELISA result, 95 of the 102 (93.1%) with a self-reported prior positive test result were positive for SARS-CoV-2 IgG antibodies. Of the 582 returning students with ELISA results who did not report a positive SARS-CoV-2 test, 113 (16.5%) were positive for SARS-CoV-2 IgG antibodies. Of the total 684 returning students with ELISA results, 208 (30.41%) were positive for SARS-CoV-2 IgG antibodies (Figure 2). Among the community resident participants, 42 of 1313 (3.2%) were positive for SARS-CoV-2 antibodies at their first visit (Figure 2). Between their first and second visit, 54 participants converted from negative to positive and 19 converted from positive to negative; 96 (7.3%) were positive for SARS-CoV-2 IgG antibodies at either visit.

**Table 3:**
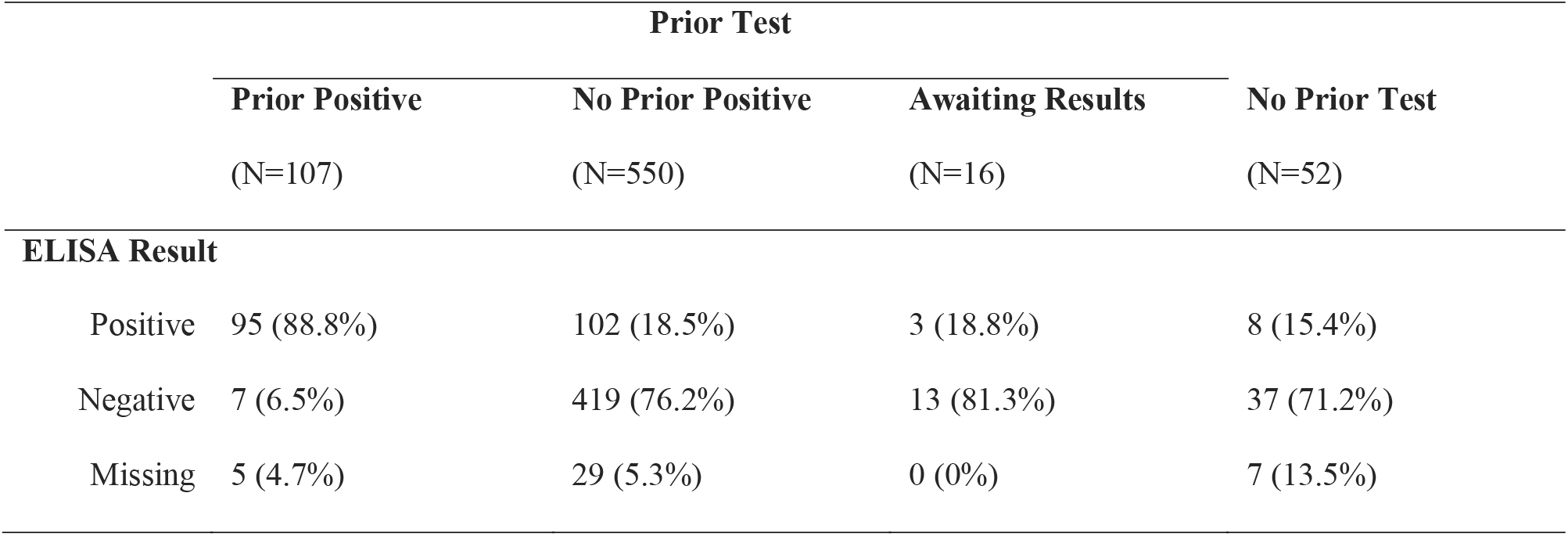
IgG ELISA results as a function of self-reported prior SARS-CoV-2 diagnostic test outcome among returning student cohort participants.

**Figure 2:**
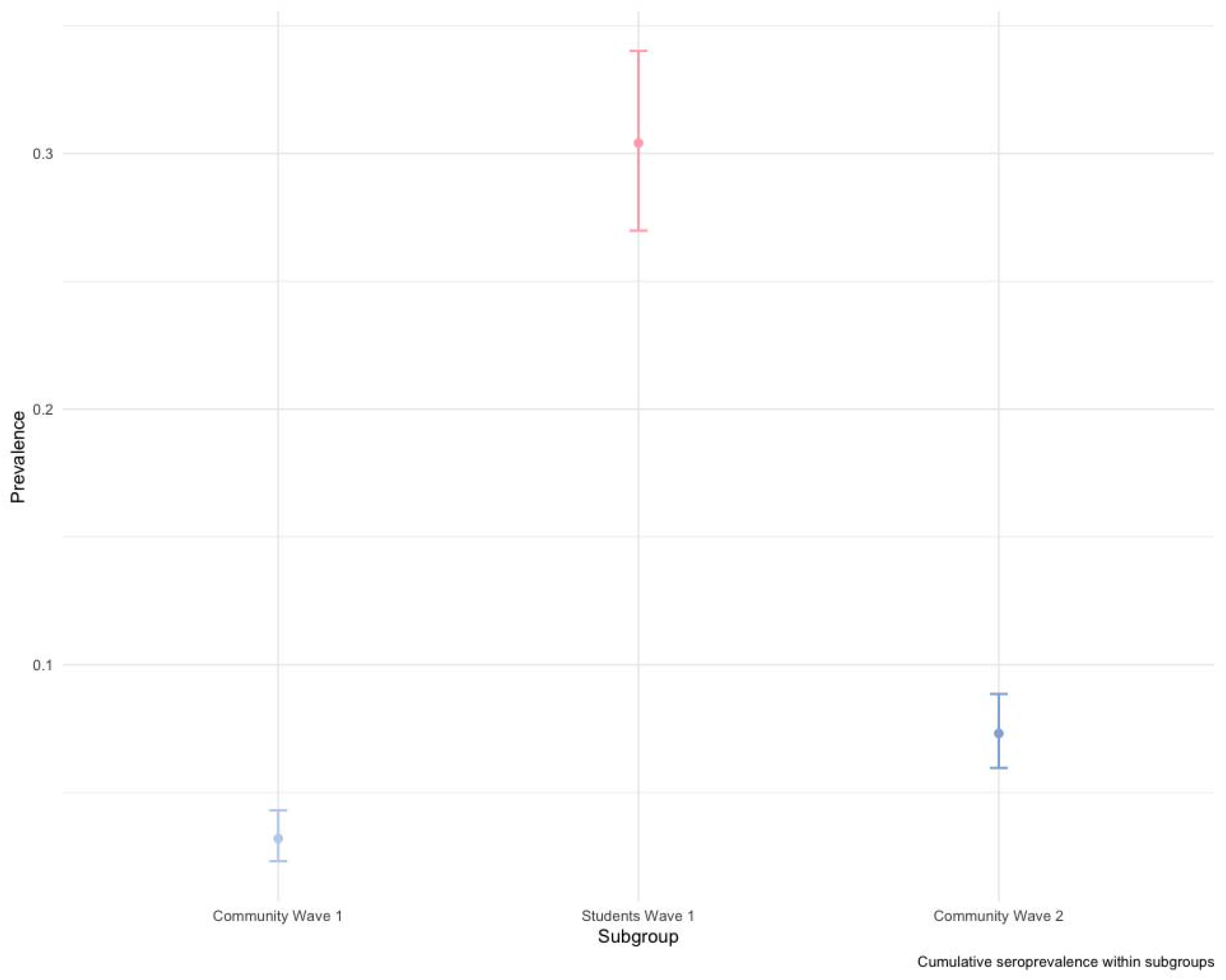
Raw seroprevalence (circles) with 95% binomial confidence intervals for the community residents at the first visit at the start of the Fall 2020 term (light blue), returning students at the end of the fall 2020 term (red), and community residents at either the first or the second visit after student departure (dark blue).

Of the returning students with a self-reported prior positive SARS-CoV-2 test, 93.1% (95% CrI: 86.4-97.2%) had positive IgG antibodies; this was used as an estimate of sensitivity of the IgG assay for detecting previously detectable infection (see Supplement for an alternative calculation of sensitivity that includes community resident responses). For all values of specificity below 0.95, the 95% credible intervals for the prevalence in the community residents overlapped for the pre- and post-term time points, and neither overlapped with the returning student subgroup (Figure 3).

**Figure 3:**
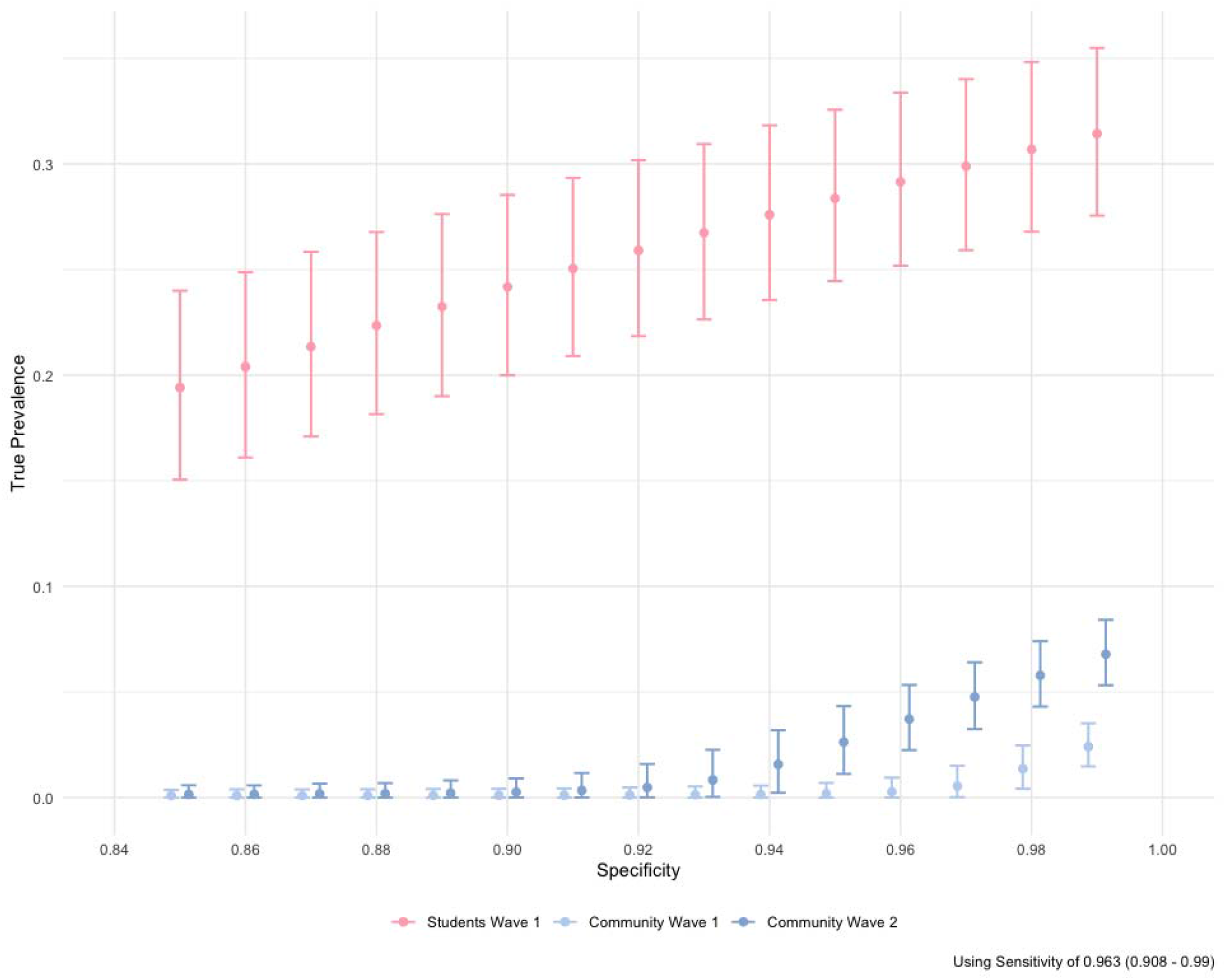
Estimated true prevalence (circles, with 95% confidence intervals) among participants at each sampling interval corrected for estimated assay sensitivity as a function of the assumed assay specificity (x-axis). Light blue indicates community residents at the first visit at the start of the Fall 2020 term, red indicates returning students at the end of the Fall 2020 term, and dark blue indicates community residents at the second visit after student departure.

Among the returning students, only close proximity to a known SARS-CoV-2-positive individual (aOR: 3.1, 2.07-4.64) and attending small gatherings in the past 3 months (aOR: 1.52, 1.03-2.24) were significantly associated with a positive ELISA classification in the multivariable model (Table 4). Attending medium gatherings (OR: 1.78, 1.17-2.69), and close proximity to an individual showing key COVID-19 symptoms (OR: 1.67, 1.19-2.36) were also associated with the IgG positivity in crude calculations of association. Among the community cohort, the amount of student contact was not associated with cumulative IgG positivity. However, PSU employees experienced reduced odds of positivity (OR: 0.56, 0.35-0.9). Neither AIC or BIC were improved by the addition of student contact as a variable over employment status only, or using student contact as the only variable.

**Table 4:** Crude and adjusted odds ratios (aOR) of risk factors among returning PSU UP student cohort

| Dependent: Assay |  | Negative | Positive | OR (univariable) | OR (multiple imputation) |
| --- | --- | --- | --- | --- | --- |
| Close proximity to known COVID-19 Positive Individual | No | 277 (58.3%) | 61 (29.5%) | • | • |
|  | Yes | 198 (41.7%) | 146 (70.5%) | 3.35 (2.37-4.78, p<0.001) | 3.10 (2.07-4.64, p<0.001) |
| Close proximity to individual showing COVID-19 symptoms | No | 346 (73.0%) | 128 (61.8%) | • | • |
|  | Yes | 128 (27.0%) | 79 (38.2%) | 1.67 (1.18-2.36, p=0.004) | 0.87 (0.58-1.30, p=0.494) |
| Travelled in the 3 months prior to campus arrival | No | 209 (45.4%) | 82 (40.8%) | • | • |
|  | Yes | 251 (54.6%) | 119 (59.2%) | 1.21 (0.86-1.69, p=0.269) | 1.05 (0.73-1.53, p=0.785) |
| Travelled since campus arrival | No | 183 (38.5%) | 82 (39.6%) | • | • |
|  | Yes | 292 (61.5%) | 125 (60.4%) | 0.96 (0.68-1.34, p=0.789) | 0.85 (0.59-1.23, p=0.394) |
| Attended a gathering of 20-50 people since arrival for the Fall Semester | No | 280 (59.1%) | 82 (39.6%) | • | • |
|  | Yes | 194 (40.9%) | 125 (60.4%) | 2.20 (1.58-3.08, p<0.001) | 1.52 (1.03-2.24, p=0.034) |
| Attended a gathering of 51-1000 people since arrival for the Fall Semester | No | 396 (85.3%) | 154 (76.6%) | • | • |
|  | Yes | 68 (14.7%) | 47 (23.4%) | 1.78 (1.17-2.69, p=0.007) | 1.32 (0.83-2.10, p=0.238) |
| Ate in a dining hall in the past 7 days | No | 394 (83.1%) | 163 (79.1%) | • | • |
|  | Yes | 80 (16.9%) | 43 (20.9%) | 1.30 (0.85-1.96, p=0.214) | 1.30 (0.74-2.28, p=0.356) |
| Ate in a restaurant in the past 7 days | No | 250 (52.5%) | 96 (46.8%) | • | • |
|  | Yes | 226 (47.5%) | 109 (53.2%) | 1.26 (0.91-1.75, p=0.173) | 1.12 (0.78-1.61, p=0.539) |
| Only ate in their room in the past 7 days | No | 158 (33.2%) | 76 (36.9%) | • | • |
|  | Yes | 318 (66.8%) | 130 (63.1%) | 0.85 (0.61-1.20, p=0.350) | 0.91 (0.61-1.34, p=0.625) |
| Lives in University housing | No | 349 (73.5%) | 152 (73.4%) | • | • |
|  | Yes | 126 (26.5%) | 55 (26.6%) | 1.00 (0.69-1.45, p=0.991) | 0.89 (0.54-1.45, p=0.630) |

Both the returning students and community residents self-reported high masking compliance; 86.7% and 75.9%, respectively, reported always wearing mask or cloth face covering when in public (Table S1, Table S2). Less than one third of both groups (28.9% and 29.8%, respectively) self-reported always maintaining 6-feet of distance from others in public. Less than half (42.8%) of returning students indicated that they always avoided groups of 25 or greater, in contrast with 65.7% of community residents.

## Discussion

The return of students to in-person instruction on the PSU UP campus was associated with a large increase in COVID-19 incidence in the county, evidenced by over 4,500 student cases at PSU [18]. In a sample of 684 returning students, 30.4% were positive for SARS-CoV-2 antibodies. Out of approximately 35,000 students who returned to campus, this implies that the detected cases may account for ∼40% of all infections among PSU UP students. Despite this high overall incidence of SARS-CoV-2 infection in the county during the Fall 2020 term, the studied cohort of community residents (who disproportionately identified as female and lived in close proximity to campus) saw only a modest increase in the prevalence of SARS-CoV-2 IgG antibodies (3.2 to 7.3%) between September and December 2020; consistent with a nation-wide estimate of seroprevalence for the summer of 2020 [23]. The true prevalence of prior SARS-CoV-2 infection in the cohorts depends on the assumed sensitivity and specificity. However, for most realistic values of sensitivity and specificity there was little evidence of a significant increase among the community resident sample. While in-person student instruction has been associated with an increase in per-capita COVID-19 incidence [12], these results suggest that outbreaks in the returning student and community resident cohorts we studied were asynchronous, implying limited between-cohort transmission. A recent analysis of age-specific movement and transmission patterns in the US suggested that individuals between the ages of 20-34 disproportionately contributed to spread of SARS-CoV-2 [28]. Despite close geographic proximity to a college-aged population, transmission in our community resident sample appears distinctly lagged; suggestive of the potential for health behaviours to prevent infection.

Within the student group, presence of SARS-CoV-2 antibodies was significantly associated with close proximity to known SARS-CoV-2-positive individuals and attendance of small events. No other risk factors were correlated with an increase in IgG test positivity, aligning with other research [23]. It is not possible to discern how much the likelihood of contact with a SARS-CoV-2 positive individual is due to the high campus prevalence versus individual behaviours.

Considered independently, eating in dining halls within the past 7 days was weakly associated with testing positive for SARS-CoV-2 antibodies, and participation in medium-sized events (51-1000 individuals) and close proximity to a symptomatic individual were significantly associated, which is consistent with patterns observed elsewhere [26,27]. Within the community group, being a PSU employee was significantly associated with lower odds of IgG positivity. There were no significant differences in the age distributions by employment status. Bharti *et al*. [29] identified lower per-capita incidence in Centre County residents relative to the 5 surrounding counties, as well as a greater movement restriction and less time spent outside the home. Whilst this paper only examined Centre County residents, it is plausible that PSU employees were more able to work remotely and similarly reduced their movement and non-household contacts, relative to non-PSU employees. The low number of positive community cases meant that it was not possible to identify other associations with IgG positivity.

Neither the resident nor the student participants were selected using a probability-based sample. Thus, these participants may not be representative of the populations. Those who chose to participate in this study may have been more cognizant and compliant with public health mitigation measures. Specifically, the resident participants disproportionately lived in the townships immediately surrounding the UP campus, where extensive health messaging [30] and preventative campaigns were enacted, and they have a higher median income than the residents of Centre County overall.

Though the participants reflect a convenience sample, the large differences in SARS-CoV-2 seroprevalence suggest that the cohorts did not experience a synchronous, well-mixed epidemic despite their close geographic proximity. College campuses have been observed to have high COVID-19 attack rates, and counties containing colleges and universities have been observed to have significantly higher COVID-19 incidence than demographically matched counties without such institutions [12]. Thus, while college and university operations may present a significant exposure risk, this analysis suggests the possibility that local-scale heterogeneity in mixing may allow for asynchronous transmission dynamics despite close geographic proximity. Thus, the disproportionately high incidence in the student population, which comprises less than one quarter of the county population, may bias assessment of risk in the non-student population. Risk assessment in spatial units (e.g., counties) that have strong population sub-structuring should consider these heterogeneities and their consequences to inform policy.

While SARS-CoV-2 transmission between the student and community resident populations is likely to have occurred (perhaps multiple times), the large difference in seroprevalence between the student and resident participants after the Fall term are consistent with either rare or non-persistent transmission events between students and residents, or both. This suggests that it is possible to minimize risks brought about by sub-populations with high SARS-CoV-2 incidence using behavioural interventions. This observation may have implications for outbreak management in other high risk, highly mobile populations (e.g., displaced populations, seasonal workers, military deployment). However, we note that this was achieved in the context of disproportionate investment in prevention education, testing, contact tracing, and infrastructure for isolation and quarantine by PSU in the high-prevalence sub-population (students).

With respect to the health behaviours measured, both students and community residents reported high masking rates (>75%) and low distancing rates in public (<30%). However, students had significantly higher masking and gathering rates than community residents, thus a next step is to identify factors that may explain these differences. Minimizing risk, however, may come at significant social, psychological, educational, economic, and societal costs [31]. Thus, operational planning for both institutions of higher education and their resident communities should consider both the risk of SARS-CoV-2 transmission and the costs of mitigation efforts.

## Data Availability

At present, data is deemed to contain personal health information (PHI) so cannot be shared as per the IRB approval. However, at a later date, a clean dataset without PHI will be added to a public GitHub repository, and the submission information will be updated to include the appropriate links to the data and analysis files.

## Author Contributions

*Conceptualization:* MJF, NB, MS, AR, VK

*Data curation:* MJF, CA

*Formal analysis:* CA, MJF, CMH

*Funding acquisition:* MJF, AR

*Investigation:* SS, AG, MMS, CJR

*Methodology:* CA, MJF

*Project administration:* MJF, MMS

*Software:* CA, MJF, CMH

*Supervision:* MJF, VK, SK

*Validation:* CA, MJF, CMH, SS

*Visualization:* CA, MJF

*Writing - original draft:* CA, SS, CMH, VK, MJF

*Writing - review and editing:* all authors.

## Data Access, Responsibility, and Analysis

Callum Arnold and Dr. Matthew J. Ferrari had full access to all the data in the study and take responsibility for the integrity of the data and the accuracy of the data analysis. Callum Arnold, Dr. Matthew J. Ferrari (Department of Biology, Pennsylvania State University), and Dr. Catherine M. Herzog (Huck Institutes of the Life Sciences, Pennsylvania State University) conducted the data analysis.

## Collaborators

1. Florian Krammer, Mount Sinai, USA for generously providing the transfection plasmid pCAGGS-RBD
2. Scott E. Lindner, Allen M. Minns, Randall Rossi produced and purified RBD
3. The D4A Research Group: Dee Bagshaw, Clinical & Translational Science Institute, Cyndi Flanagan, Clinical Research Center and the Clinical & Translational Science Institute, Thomas Gates, Social Science Research Institute, Margeaux Gray, Dept. of Biobehavioral Health, Stephanie Lanza, Dept. of Biobehavioral Health and Prevention Research Center, James Marden, Dept. of Biology and Huck Institutes of the Life Sciences, Susan McHale, Dept. of Human Development and Family Studies and the Social Science Research Institute, Glenda Palmer, Social Science Research Institute, Rachel Smith, Dept. of Communication Arts and Sciences and Huck Institutes of the Life Sciences, and Charima Young, Penn State Office of Government and Community Relations.
4. The authors thank the following for their assistance in the lab: Liz D. Cambron, Elizabeth M. Schwartz, Devin F. Morrison, Julia Fecko, Brian Dawson, Sean Gullette, Sara Neering, Mark Signs, Nigel Deighton, Janhayi Damani, Mario Novelo, Diego Hernandez, Ester Oh, Chauncy Hinshaw, B. Joanne Power, James McGee, Riëtte van Biljon, Andrew Stephenson, Alexis Pino, Nick Heller, Rose Ni, Eleanor Jenkins, Julia Yu, Mackenzie Doyle, Alana Stracuzzi, Brielle Bellow, Abriana Cain, Jaime Farrell, Megan Kostek, Amelia Zazzera, Sara Ann Malinchak, Alex Small, Sam DeMatte, Elizabeth Morrow, Ty Somberger, Haylea Debolt, Kyle Albert, Corey Price, Nazmiye Celik

