## Supplemental text/tables/figures for "SARS-CoV-2 Seroprevalence in a University Community: A Longitudinal Study of the Impact of Student Return to Campus on Infection Risk Among Community Members"

Supplemental Appendix

### Supplementary Text

#### Laboratory Methods:

##### Production and Purification of SARS-CoV-2 Receptor-binding domain (S/RBD)

Transfections of plasmid pSL1510 (pCAGGS-RBD from Florian Krammer, Mount Sinai, USA) was performed using the Expi293 Expression System from ThermoFisher. Cells were cultured per manufacturer’s instructions (37°C, 8% CO2, in shaker flasks at 120-130 rpm), and the supernatant was harvested by simple centrifugation on the third day for downstream processing. Cell viability and concentration were monitored throughout to ensure that the culture remained in log phase growth. The detailed protocol is deposited in protocols.io [1]. Briefly, culture supernatant was incubated with pre-equilibrated Ni-NTA resin in 1X PBS at 4°C for 1 h on a nutator, after which a gravity column was used to elute the protein.

##### Estimation of IgG antibodies against SARS-CoV-2

An in-house indirect isotype-specific (IgG) ELISA against SARS-CoV-2 receptor-binding domain (S/RBD) was developed [1]. Commercially purchased human monoclonal antibody reactive to spike regions of SARS-CoV-1 and SARS-CoV-2 were used as positive controls in the assay (Two isotypes of CR3022, IgG1: Ab01680-10.0; Absolute Antibody, USA). The cut-off for this IgG ELISA was determined as an optical density (absorbance at 450 nm) higher than three or six standard deviations above the mean of the tested pre-COVID-19 serum samples (n=100). Briefly, serum was separated from the blood collected from study participants and inactivated at 56°C for 30 minutes. Microtiter plates were coated with purified recombinant S/RBD. Negative serum control was included on each microtiter plate. 1:50 dilutions of serum were added, incubated for 1 hour, washed, incubated with goat anti-human IgG (Fc specific) (A0170, Sigma-Aldrich, USA), and washed. 3,3′,5,5′-Tetramethylbenzidine dihydrochloride (TMB) was used as the ELISA substrate (T3405, Sigma-Aldrich, USA) was added, the plates were developed until the top dilution reached the saturation pointes, and the reaction was stopped with H2SO4. Plates were read at an absorbance of 450 nm.

#### Statistical Methods

##### Treatment of Missing Data

In the subset of individuals in the returning student subgroup that had ELISA results, there are few missing values for the model variables, with the exception of “working as a service professional” (421/684). As a result of high missingness, service professional was removed as a predictor in the model. Exploration of the missing values in the remaining predictor variables demonstrate no bias by outcome, confirmed using Chi-squared tests of missingness in predictors by outcome level. Little’s test of Missing Completely At Random (MCAR) indicated that the data was not MCAR (p = 0.0727533)[2], and three imputation methods (MICE, k-Nearest Neighbour with 5 neighbours, and Bagged Tree) [3–5] were used to compare model fits (Supplemental Figure 1). Most missing values occurred across all variables, and there was no observable pattern among the majority of variables: there was some evidence that missingness in “travel in the 3 months prior to return” was associated with “travelling since campus return” response, and that missingness in “eaten in a restaurant in the past 7 days” was associated with “IgG classification.” As such, the predictor variables were deemed to be ‘Missing At Random,’ and MICE was used to impute missing values.

##### Alternative Estimate of True Prevalence

In the main text we present estimates of the true prevalence in the returning student and community resident cohorts that corrects for the sensitivity of the assay. We estimated sensitivity based on the returning student samples only because the student population had high access to RT-PCR diagnostic tests. Here we present an alternative analysis using an estimate of sensitivity including the community residents. 9 community residents self-reported a positive COVID-19 diagnosis by a medical professional prior to the first visit; an additional 19 community residents reported a positive COVID-19 diagnosis between the first and second visit. Of these, 17 were positive for IgG antibodies. Pooled with the student results, this results in a sensitivity estimate of 89 (95% CI: 82.5-93.7). This implies a lower sensitivity in the community resident participants, though the number of observations is low. Supplemental Figure 2 shows the estimated true prevalence assuming a uniform prior on the interval (82.5, 93.7) on sensitivity in the community resident population and a uniform prior on the interval (0.85, 0.99) on specificity. For all values of specificity greater than 0.85, there is no change in the qualitative result that the 95% confidence intervals for prevalence in the community residents overlap for both visits for specificity values less than 0.95, and are distinctly different to the prevalence within the returning student subgroup.

### Tables

Supplemental Table 1: Propensity of following public health measures in returning students and community members with PSU ELISA results; subset of community members that received the “Health Messaging” survey. P-value refers to Chi-square test with Yates’ continuity correction of proportions in the predictor level by cohort.

| **PH Measure** |  | **Community - Health Messaging** | **Returning Students** | **p** |
| --- | --- | --- | --- | --- |
| Total N (%) |  | 835 (55.0%) | 684 (45.0%) |  |
| Mask Wearing | Always | 633 (76.1%) | 593 (87.0%) | <0.001 |
|  | Not Always | 199 (23.9%) | 89 (13.0%) |  |
| Distancing in Public | Always | 249 (30.0%) | 198 (29.1%) | 0.749 |
|  | Not Always | 582 (70.0%) | 483 (70.9%) |  |
| Avoiding crowds of >25 people | Always | 549 (65.8%) | 293 (43.0%) | <0.001 |
|  | Not Always | 285 (34.2%) | 389 (57.0%) |  |

Supplemental Table 2: Raw prevalence in each subgroup by adherence to PH public health measures

|  | **Community - Health Messaging** | | **Returning Students** | |
| --- | --- | --- | --- | --- |
|  | **Negative** | **Positive** | **Negative** | **Positive** |
|  | (N=804) | (N=31) | (N=476) | (N=208) |
| **Mask Wearing** | | | | |
| Always | 610 (75.9%) | 23 (74.2%) | 410 (86.1%) | 183 (88.0%) |
| Not Always | 191 (23.8%) | 8 (25.8%) | 65 (13.7%) | 24 (11.5%) |
| Missing | 3 (0.4%) | 0 (0%) | 1 (0.2%) | 1 (0.5%) |
| **Distancing in Public** | | | | |
| Always | 242 (30.1%) | 7 (22.6%) | 150 (31.5%) | 48 (23.1%) |
| Not Always | 558 (69.4%) | 24 (77.4%) | 324 (68.1%) | 159 (76.4%) |
| Missing | 4 (0.5%) | 0 (0%) | 2 (0.4%) | 1 (0.5%) |
| **Avoiding crowds of >25 people** | | | | |
| Always | 530 (65.9%) | 19 (61.3%) | 219 (46.0%) | 74 (35.6%) |
| Not Always | 273 (34.0%) | 12 (38.7%) | 256 (53.8%) | 133 (63.9%) |
| Missing | 1 (0.1%) | 0 (0%) | 1 (0.2%) | 1 (0.5%) |

### Figures

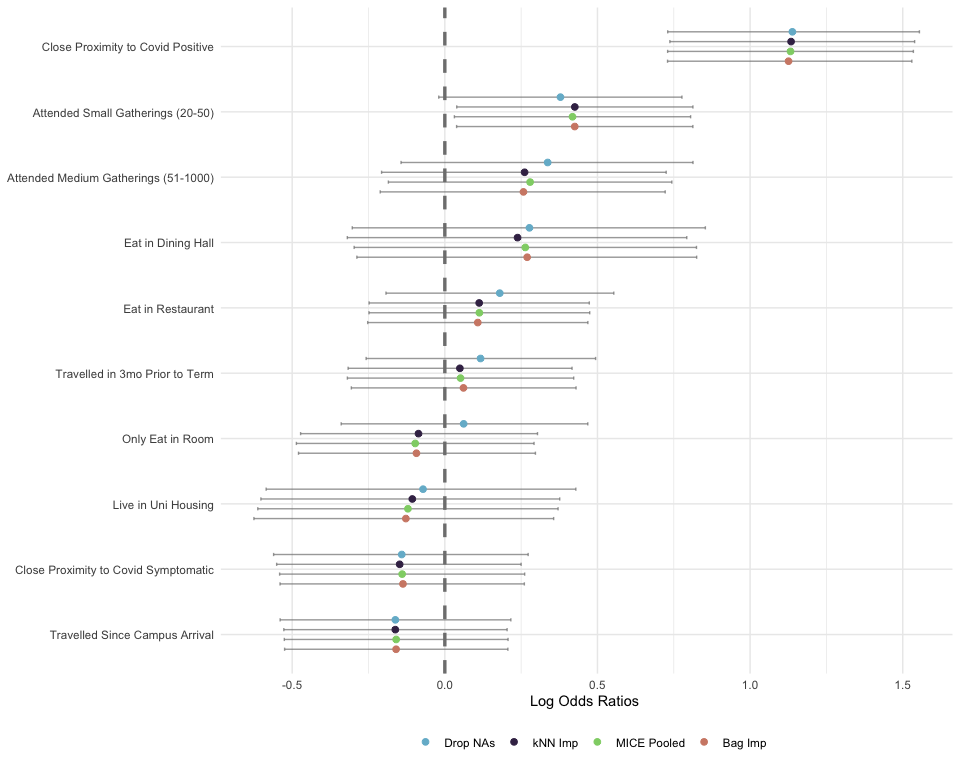

Supplemental Figure 1: Imputation method comparison among returning students

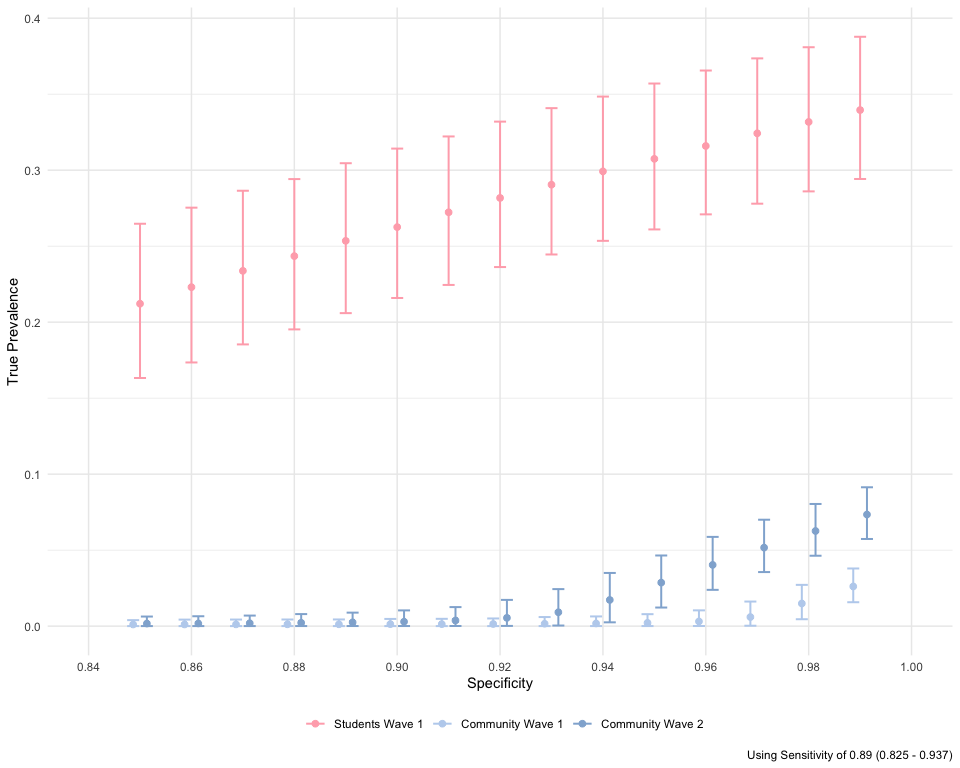

Supplemental Figure 2: Sensitivity analysis of true prevalence amongst returning student and community subgroups, using pooled estimate of IgG test sensitivity against self-reported prior positive test
